# Health and Demographic Impact on COVID-19 Infection and Mortality in US Counties

**DOI:** 10.1101/2020.05.06.20093195

**Authors:** Zidian Xie, Dongmei Li

## Abstract

**Introduction:** With the pandemic of COVID-19, the number of confirmed cases and related deaths are increasing in the US. We aimed to understand the potential impact of health and demographic factors on the infection and mortality rates of COVID-19 at the population level.

**Methods:** We collected total number of confirmed cases and deaths related to COVID-19 at the county level in the US from January 21, 2020 to April 23, 2020. We extracted health and demographic measures for each US county. Multivariable linear mixed effects models were used to investigate potential correlations of health and demographic characteristics with the infection and mortality rates of COVID-19 in US counties.

**Results:** Our models showed that several health and demographic factors were positively correlated with the infection rate of COVID-19, such as low education level and percentage of Black. In contrast, several factors, including percentage of smokers and percentage of food insecure, were negatively correlated with the infection rate of COVID-19. While the number of days since first confirmed case and the infection rate of COVID-19 were negatively correlated with the mortality rate of COVID-19, percentage of elders (65 and above) and percentage of rural were positively correlated with the mortality rate of COVID-19.

**Conclusions:** At the population level, health and demographic factors could impact the infection and mortality rates of COVID-19 in US counties.

## Introduction

Since the first reported coronavirus disease 2019 (COVID-19) case was identified in Wuhan, China ^1^, it quickly spreads out and was declared as a public health emergency of international concern on January 30, 2020 and a pandemic on March 11, 2020 by the World Health Organization (WHO). By April 28, 2020, there are nearly three millions confirmed COVID-19 cases and over 200,000 confirmed deaths globally ^2^.

Much effort has been put in understanding the COVID-19 virus infection mechanism and further developing effective vaccines for COVID-19 ^3-5^. While the COVID-19 vaccines are under development, it becomes important to determine what factors might contribute to the infection and mortality of COVID-19, which might help us slow down the pandemic of COVID-19 and reduce the economic and human losses due to COVID-19. It has been shown that travel restrictions are effective in controlling the COVID-19 epidemic in China ^6^. Racial disparities were reported to emerge with the infection and fatality of COVID-19. For example, African American are in higher risk of COVID-19 infection and fatality ^7^

While COVID-19 becomes an epidemic in the US, there are geographic variations in the number of COVID-19 confirmed cases and related deaths, which might due to the differences in epidemiologic and population-level factors ^8^. In this study, at the population level, we investigated potential correlations of health and demographic factors with the infection and mortality rates of COVID-19 in US counties.

## Methods

### Data Source

The number of confirmed COVID-19 cases and related deaths in each US counties from January 21, 2020 to April 23, 2020 were obtained from the website of “1Point3Acres.com” (https://coronavirus.1point3acres.com). Some county-level information (such as the population and land area in 2018, economic typology in 2015) are downloaded from United States Census Bureau (https://www.census.gov) and USDA Economic Research Service (https://www.ers.usda.gov/data-products/county-level-data-sets/download-data/). The health-related and other demographic information at the county level are obtained from County Health Rankings & Roadmaps (CHR&R, www.countyhealthranking.org) in 2020. CHR&R is a collaborating program between the Robert Wood Johnson Foundation (RWJF) and the University of Wisconsin Population Health Institute, which provide a number of measures related to the health of nearly all counties in the US, including health behaviors, clinical care, social and economic factors, and physical environment. These measures are standardized and combined using scientifically-informed weights.

### Outcome Variables and Covariates

The outcome variables include the infection rate of COVID-19 (the number of confirmed cases divided by the population), and the mortality rate of COVID-19 (the number of deaths divided by the number of confirmed cases) in each US county. We did a log-transformation on the infection and mortality rates of COVID-19.

The covariates we considered include health and demographic measures at the county level. Health measures include: number of days since first confirmed COVID-19 case in each county, metropolitan (two levels: yes and no) in 2013, economic typology in 2015 (six levels: nonspecialized, farm-dependent, mining-dependent, manufacturing-dependent, government-dependent, and recreation), percentage of poverty in 2018, percentage of fair or poor health, average number of physically unhealthy days, average number of mentally unhealthy days, percentage of smokers, percentage of adults with obesity, percentage of physically inactive, percentage of with access to exercise opportunities, percentage of excessive drinking, percentage of uninsured, percentage of vaccinated, percentage of unemployed, average daily PM2.5, presence of water violation (two levels: yes and no), percentage of frequent physical distress, percentage of frequent mental distress, percentage of adults with diabetes, percentage of food insecure (percentage of population who lack adequate access to food), percentage of insufficient sleep, median household income, segregation index (the degree to which the minority group is distributed differently than whites across census tracts), percentage of homeowners, percentage of severe housing cost burden (percentage of households that spend 50% or more of their household income on housing). Demographic factors include: percentage of less than 18 years of age, percentage of 65 and over, percentage of Black, percentage of American Indian & Alaska Native, percentage of Asian, percentage of Native Hawaiian/Other Pacific Islander, percentage of Hispanic, percentage of Non-Hispanic White, percentage of not proficient in English, percentage of female, percentage of rural, percentage of adults with a high school or less education, population in 2018, population density (# people per square miles).

### Statistical Analysis

Multivariable linear mixed effects models were used to investigate the correlations between the outcome variables and prediction variables. The correlations of counties within the same state were considered through the compound symmetry variance-covariance structure, which assumes the correlations of counties are the same within the same state. Covariates controlled for in our final statistical models were selected through purposeful selection of covariates method ^9^. The coefficient estimates and their 95% confidence intervals (CIs) were used to quantify the correlation. Positive estimate indicates positive correlation while negative estimate indicates negative correlation between covariates and outcome variables. P-value below 0.05 indicates a significance contribution of a variable to the statistical models. All analyses were conducted using proc mixed procedure in SAS V.9.4 (SAS Institute Inc., Cary, NC).

## Results

### Health and Demographic Impact on the Infection Rate of COVID-19

Among 3,143 counties and county equivalents of the United States, by April 23, 2020, based on the website of 1Point3Acres.com, there were 2,743 US counties that had reported COVID-19 cases, ranging from 1 to 147,297 cases with total confirmed cases of 855,979. Considering the difference in total population among different counties, we calculated the COVID-19 infection rate as the number of COVID-19 cases divided by the total population in each county. To investigate the potential correlation of health and demographic factors with the COVID-19 infection rate at the county level, we performed a multivariable linear mixed effects model.

As shown in Table 1, a number of health and demographic factors showed positive correlations with the infection rate of COVID-19. With the increase of the number of days since the first reported case in each county, the total infection rate of COVID-19 in log scale increased (b = 0.02175, 95% CI: 0.01600, 0.02750). The higher percentage of adults with a high school or less education, the higher of COVID-19 infection rate in log scale (b = 0.02113, 95% CI: 0.01491, 0.02734). The population density (the number of people per square miles) was positively correlated with COVID-19 infection rate in log scale although the estimated coefficient is very small (b = 0.00004, 95% CI: 0.00002, 0.00006). With the higher housing cost burden, the COVID-19 infection rate in log scale significantly increased (b = 0.03451, 95% CI: 0.01786, 0.05116). With the higher percentage of Black or American Indian & Alaska Native, the COVID-19 infection rate in log scale increased significantly. The COVID-19 infection rate in log scale was also positively correlated with percentage of people who are not proficient in English (b = 0.05180, 95% CI: 0.02359, 0.08001). In addition, a number of health and demographic factors showed negatively correlations with the infection rate of COVID-19 in log scale (Table 1). With higher percentage of smokers, the COVID-19 infection rate in log scale decreased significantly (b = −0.03747, 95% CI: −0.06610, −0.00884). With the increase in percentage of adults with obesity, excessive drinking and food insecure, the infection rate of COVID-19 in log scale decreased significantly. With the higher of segregation index, the lower of COVID-19 infection rate in log scale (b = −0.00457, 95% CI: −0.00723, −0.00192). The percentage of Hispanic in the population showed a negative correlation with the infection rate of COVID-19 in log scale (b = −0.01236, 95% CI: −0.01957, −0.00515).

**Table 1.** The Impact of Health and Demographic Factors on COVID-19 Infection Rate (in Log Scale) in US Counties.

| **Variables** | **Estimated Coefficients (b)** | **Standard Error** | **P-value** | **Confidence Interval** |
| --- | --- | --- | --- | --- |
| **Days since first case** | 0.04476 | 0.00198 | <.0001 | (0.04087, 0.04865) |
| **Percentage of adults with a high school or less education** | 0.02113 | 0.00317 | <.0001 | (0.01491, 0.02734) |
| **Population density** | 0.00004 | 0.00001 | <.0001 | (0.00002, 0.00006) |
| **Percentage of severe housing cost burden** | 0.03451 | 0.00849 | <.0001 | (0.01786, 0.05116) |
| **Percentage of Black** | 0.03394 | 0.00263 | <.0001 | (0.02877, 0.03910) |
| **Percentage of American Indian & Alaska Native** | 0.03443 | 0.00532 | <.0001 | (0.02399, 0.04486) |
| **Percentage of people not proficient in English** | 0.05180 | 0.01438 | 0.0003 | (0.02359, 0.08001) |
| **Percentage of smokers** | -0.03747 | 0.01459 | 0.0104 | (-0.06610, -0.00884) |
| **Percentage of adults with obesity** | -0.01133 | 0.00441 | 0.0102 | (-0.01998, -0.00269) |
| **Percentage of excessive drinking** | -0.02717 | 0.01196 | 0.0232 | (-0.05062, -0.00371) |
| **Percentage of food insecure** | -0.08426 | 0.01353 | <.0001 | (-0.11080, -0.05772) |
| **Segregation index** | -0.00457 | 0.00135 | 0.0007 | (-0.00723, -0.00192) |
| **Percentage of Hispanic** | -0.01236 | 0.00368 | 0.0008 | (-0.01957, -0.00515) |

### Health and Demographic Impact on the Mortality Rate of COVID-19

While the COVID-19 infection rates were different among US counties, the COVID-19 mortality rate (the number of COVID-19 related deaths divided by the number of confirmed cases) also varied among US counties, ranging from 0 to 1 with a mean of 0.036 (standard deviation = 0.079). To examine the impact of health and demographic factors on the mortality rate of COVID-19 in US counties, a multivariable linear mixed effects model was performed. As shown in Table 2, several health and demographic factors were significantly negatively correlated with the COVID-19 mortality rate in log scale at the population level. With the number of days since the first confirmed COVID-19 case increased, the mortality rate in log scale significantly decreased (b = −0.02041, 95% CI: −0.02612, −0.01470). The COVID-19 infection rate was significantly negatively correlated with the mortality rate in log scale (b = −44.12260, 95% CI: −58.21640, −30.02880). Higher percentage of vaccination significantly correlated with lower COVID-19 mortality rate in log scale (b = −0.01153, 95% CI: −0.01860, −0.00446). In contrast, the percentage of elders aged 65 and over was significantly positively correlated with the COVID-19 mortality rate in log scale (b = 0.04734, 95% CI: 0.03509, 0.05958). The percentage of rural was positively correlated with the mortality rate of COVID-19 in log scale (b = 0.00322, 95% CI: 0.00116, 0.00529). The correlation of population density with the COVID-19 mortality rate in log scale was positive and significant although the estimated coefficient is small (b = 0.00007, 95% CI: 0.00005, 0.00010). Compared to the counties with economic typology of “Recreation”, the counties with “Nonspecialized”, “Farm-dependent” or “Mining-dependent” had significantly higher COVID-19 mortality rate in log scale.

**Table 2.**
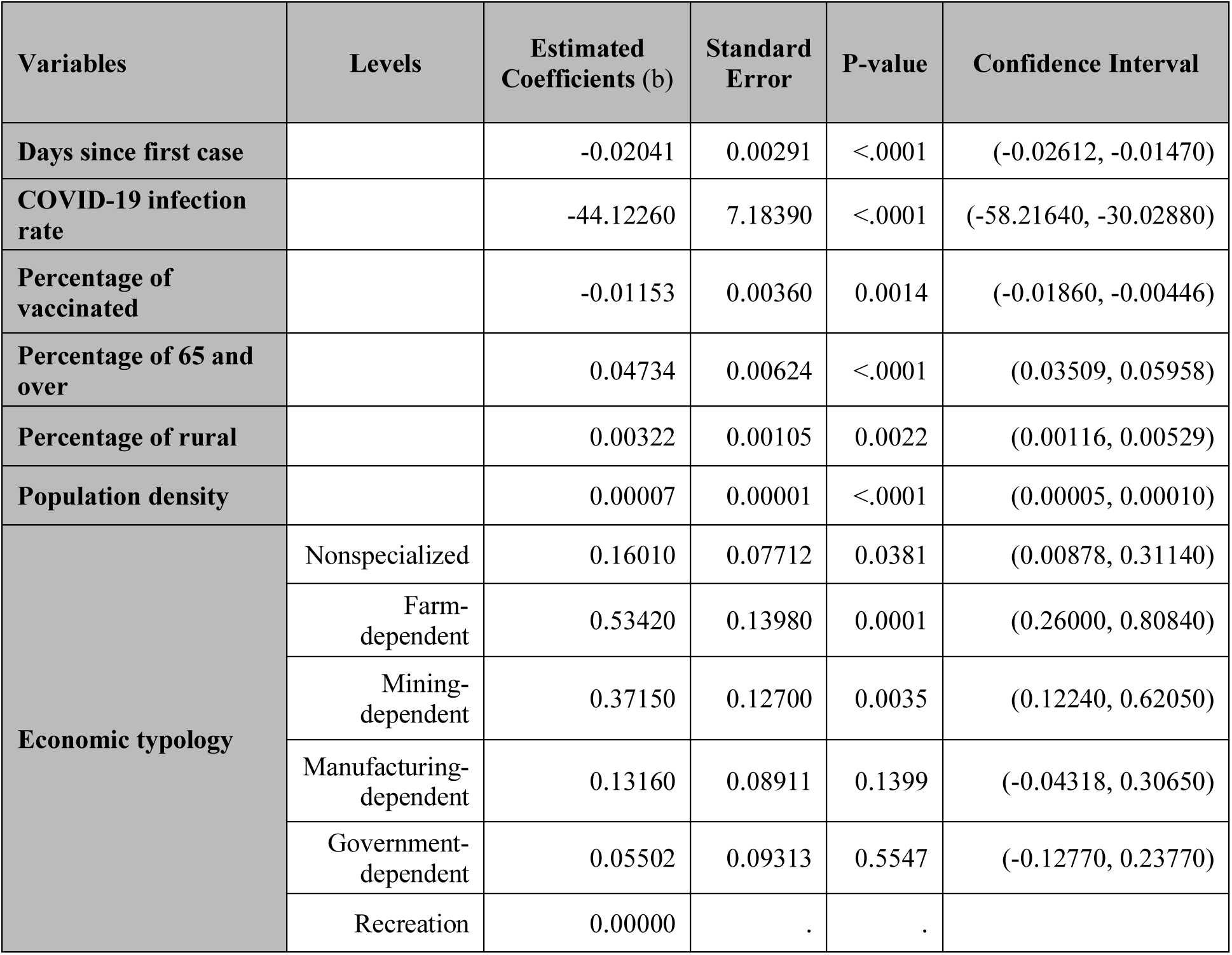
The Impact of Health and Demographic Factors on COVID-19 Mortality Rate (in Log Scale) in US Counties.

| Variables | Levels | Estimated Coefficients (b) | Standard Error | P-value | Confidence Interval |
| --- | --- | --- | --- | --- | --- |
| Days since first case |  | -0.02041 | 0.00291 | <.0001 | (-0.02612, -0.01470) |
| COVID-19 infection rate |  | -44.12260 | 7.18390 | <.0001 | (-58.21640, -30.02880) |
| Percentage of vaccinated |  | -0.01153 | 0.00360 | 0.0014 | (-0.01860, -0.00446) |
| Percentage of 65 and over |  | 0.04734 | 0.00624 | <.0001 | (0.03509, 0.05958) |
| Percentage of rural |  | 0.00322 | 0.00105 | 0.0022 | (0.00116, 0.00529) |
| Population density |  | 0.00007 | 0.00001 | <.0001 | (0.00005, 0.00010) |
| Economic typology | Nonspecialized | 0.16010 | 0.07712 | 0.0381 | (0.00878, 0.31140) |
|  | Farm-dependent | 0.53420 | 0.13980 | 0.0001 | (0.26000, 0.80840) |
|  | Mining-dependent | 0.37150 | 0.12700 | 0.0035 | (0.12240, 0.62050) |
|  | Manufacturing-dependent | 0.13160 | 0.08911 | 0.1399 | (-0.04318, 0.30650) |
|  | Government-dependent | 0.05502 | 0.09313 | 0.5547 | (-0.12770, 0.23770) |
|  | Recreation | 0.00000 | . | . |  |

## Discussions

With the pandemic of COVID-19, especially in the US, while it is of upmost importance to discover the underlying biological mechanisms of COVID-19 infection and further develop the effective vaccines and medication, it is important to understand what other factors can affect the infection and mortality of COVID-19. Here, at the population level, we investigated potential correlations of health and demographic factors with the infection and mortality rates of COVID-19 in US counties. Our statistical models showed that some health and demographic factors were positively correlated with the infection and mortality rates of COVID-19 while others showed negative correlations. For example, the number of days since the first reported case in US counties was positively correlated with the infection rate but percentage of smokers was negatively correlated with the infection rate of COVID-19. While the infection rate of COVID-19 was negatively correlated with the mortality rate, percentage of elders (65 and over) was positively correlated with the COVID-19 mortality rate.

Our results showed that while the number of days since the first reported case in each county was positively correlated with the infection rate of COVID-19, it was negatively correlated with the mortality rate of COVID-19. In addition, the infection rate was significantly negatively correlated with the mortality rate of COVID-19. One possible explanation could be that health professionals or medical system developed more efficient medical practices (such as increase ICU beds and enhance social distancing) to treat COVID-19 patients, so that the mortality rate was decreasing even though the infection rate was increasing with the spreading of COVID-19 over time.

In this study, we showed that the population density was positively correlated with both the infection and mortality rates of COVID-19 even though the estimated coefficients were small. With the higher population density, social distancing might be more challenging, which could contribute to higher infection and mortality rates of COVID-19.

Percentage of Black or American Indian & Alaska Native, as well as people that are not proficient in English was positively correlated with the infection rate of COVID-19, suggesting that these minorities might have high risk of COVID-19 infection. The Centers for Disease Control and Prevention (CDC) reported that 30% of COVID-19 patients are African American even though only 13% of the US population are African Americans ^10^. For people in racial and ethnic minority groups, their living conditions (densely populated areas due to severe housing cost burden) could contribute to their high infection rate because they might have difficulties protecting themselves from COVID-19 infection and practicing social distancing.

One study based on 140 COVID-19 patients in China showed that only 1.4% of patients were current smokers ^11^, and another study showed that 12.6% of COVID-19 patients in China were smokers ^12^, both are significantly lower than 27.3% of smokers in China. A study on 343 COVID-19 patients in France showed 5.3% were daily smokers, which was below 25.4% smoking rate in the French population ^13^. Here, we showed, at the population level, percentage of smokers in US counties was negative correlated with the infection rate of COVID-19. However, several studies showed that smoking could worsen the health outcomes of COVID-19 patients ^14,15^. While it requires further investigation on how smoking affects the infection and mortality of COVID-19, careful caution should be taken to interpret these results and smoking should be discouraged.

In this study, we showed that percentage of population with 65 and over was positively correlated with the mortality rate of COVID-19 at the population level. One study on COVID-19 patients from China and other countries showed that the older age group (above 60 years) had significantly higher death rate than the younger age group (below 60 years) ^16^. The Centers for Disease Control and Prevention reported that 8 out of 10 deaths in the US related to COVID-19 have been in adults with age 65 and over. One possible reason for high death rate in elder patients is that the majority of elder patients with COVID-19 have comorbidities, such as respiratory diseases, kidney failure and heart conditions, which might contribute to the fatal outcome or even more than the contribution of SARS-CoV-2. Therefore, the seniors need to be more careful to avoid COVID-19 infection due to their high fatality rate.

It has been shown that there was no association between influenza vaccination and coronavirus ^17^ However, one study showed that influenza vaccine derived virus interference was significantly associated with coronavirus ^18^. Here, we showed that percentage of people with vaccination was negatively correlated with the mortality rate of COVID-19. Considering the difference between influenza viruses and coronaviruses, the influenza vaccines are less likely to prevent infections with coronavirus. In this study, we did not observe the significant correlation between influenza vaccination and the infection rate of COVID-19. However, without influenza vaccination, the COVID-19 patients might have more severe symptoms with the influenza infection, which might lead to higher fatality rate. Whether influenza vaccines could lead to low mortality rate of COVID-19 requires further investigation.

Our statistical model showed that percentage of rural was positively correlated with the mortality rate of COVID-19. Moreover, the counties that are farm-dependent or mining-dependent have high mortality rate of COVID-19. One possible explanation could be that the medical systems in rural areas are not well equipped and prepared for the COVID-19 infection. This finding should be alarming since rural areas have limited medical resources to respond to this pandemic and treat COVID-19 patients ^19^. Additional measures (such as professional health experts and well-equipped hospitals) should be taken before the COVID-19 becomes epidemic in rural areas.

In our study, the number of confirmed COVID-19 cases and deaths were from 1Point3Acres.com, which is subject to some errors (such as unreported cases). In addition, we only examined the correlation of health and demographic factors with the infection and mortality rates of COVID-19 at the population level. Therefore, the causal relationship could not be established. Some important health and demographic factors, such as the prevalence of chronic diseases (asthma, diabetes, etc.) and COVID-19 testing rate, are not included in our analysis due to their unavailability. With continuous spreading of COVID-19 in the US, the number of confirmed cases and deaths at different counties are increasing, and our models based on the most-updated COVID-19 data require continuous updating. Moreover, it remains unclear why some health factors were negatively correlated with the infection rate of COVID-19, such as percentage of adults with obesity, excessive drinking or food insecure.

While our findings at the population level may require further validation using clinical data at the individual level, our study is the first one to investigate potential correlations of health and demographic factors with the infection and mortality rates of COVID-19 in US counties. While developing COVID-19 vaccines will be the most effective way to stop this COVID-19 pandemic, it is a time-consuming and expensive process ^20^. The findings from this study may help us determine the potential impact of health and demographic factors on the infection and mortality rates of COVID-19, which could facilitate us to take effective measures to slow down the pandemic of COVID-19.

## Data Availability

The number of confirmed COVID-19 cases and related deaths in each US counties were requested from the website of “1Point3Acres.com” (https://coronavirus.1point3acres.com). Health and demographic data are downloaded from United States Census Bureau (https://www.census.gov), USDA Economic Research Service (https://www.ers.usda.gov/data-products/county-level-data-sets/download-data/), and County Health Rankings & Roadmaps (CHR&R, www.countyhealthranking.org).

https://www.census.gov

https://www.ers.usda.gov/data-products/county-level-data-sets/download-data/

https://www.countyhealthranking.org

